# The Impact of Digital Technology in Care Homes on Unplanned Secondary Care Usage and Associated Costs

**DOI:** 10.1101/2023.06.13.23291324

**Authors:** Alex Garner, Jen Lewis, Simon Dixon, Nancy Jean Preston, Camila C.C.S Caiado, Barbara Hanratty, Monica Jones, Jo Knight, Suzanne M Mason

**Affiliations:** Lancaster University; University of Sheffield; Durham University; Newcastle University; University of Leeds

## Abstract

**Background:** A substantial number of emergency department (ED) attendances from care homes could be classed as avoidable. HealthCall is a technology that aims to streamline residents’ care by recording their observations electronically. Observations are fed to remote clinical staff to triage referrals. This study assessed the effectiveness of the HealthCall technology to safely reduce unplanned secondary care usage and associated costs.

**Methods:** The study involved 118 care homes across the North East from 2018-2021. Routinely collected NHS secondary care data from County Durham and Darlington NHS Foundation Trust was linked with data from the HealthCall technology App. Four outcomes were modelled monthly using Generalised Linear Mixed Models: counts of emergency attendances, emergency admissions, emergency readmissions (28-days), and length of stay of emergency admissions. A similar approach was taken for costs. The impact ofHealthCall was tested on each outcome using the models.

**Findings:** Data from 8,702 residents was used in the analysis. Results show HealthCall reduces the number of emergency attendances by 11%[6%-15%], emergency admissions by 25% 20%-39%], readmissions reduced by 29%[24%-33%] and length of stay by 11%[3%-18%] (with an additional month-by-month decrease of 28%[24%-34%]). The cost analysis found a cost reduction of £57 per resident in 2018, increasing to £113 in 2021.

**Interpretation:** The introduction of a digital technology, such as HealthCall, significantly reduces contacts with and costs resulting from unplanned secondary care usage by care home residents.

**Funding:** This work was funded by Health Data Research UK, CFC0124.

## Introduction

There are around 17,700 care homes in England with around 430,000 residents. Most residents are over 80 years old with varying levels of healthcare needs which may be complex. Hospital attendances and admissions can be hazardous for residents, with high rates of hospital-acquired infections, increased confusion, and falls.

Generally, older patients prefer to be treated at their normal place of residence, but current NHS service configurations struggle to achieve this on many occasions, despite the NHS Long Term Plan’s [1] commitment to better healthcare provision for care home residents. One of the contributory factors to this problem are high rates of emergency department (ED) attendance and hospital admissions among care home residents.

The potential scope for reducing these, and the associated patient benefits and cost savings have been explored [2], and ready access to advice from healthcare professionals was seen as being fundamental to delivering these care improvements. The development of digital technology to support shared decision making and deliver closer working between agencies may be a scalable and cost-effective method for providing timely advice and coordinated care across service boundaries. However, evidence is needed to support these hypotheses along with a clear understanding of how to implement such tools to ensure appropriate uptake.

Health Call Solutions is a digital health initiative collaboratively run by seven NHS Foundation Trusts across North East England and North Cumbria. One of the solutions provided by Health Call is the Health Call Digital Care Homes (DCH) Application (henceforth, “HealthCall”), an application (app) designed for use by staff in care homes for older people [3]. This aims to incorporate clinical expertise into decision making for the care of residents in the homes. The app provides a structured method for seeking clinical advice for the management of care home residents who become unwell. Upon implementation of the system, staff are trained to use it to record the vital signs of the residents to allow calculation of the National Early Warning Score 2 (NEWS2). Carers can also upload free text describing a resident’s condition using a Situation, Background, Assessment, Recommendation (SBAR) approach, which is a structured form of communication used to enable information to be conveyed accurately [4,5]. Information uploaded to the app is automatically fed into a Single Point of Access where clinical staff, with access to residents’ Electronic Health Records (EHRs) triage referrals and provide advice and next steps on the care for the residents.

A primary goal of the HealthCall app is to reduce avoidable secondary care for the residents in the homes, through improved access to timely clinical advice. The app replaces the traditional method of seeking advice through telephone calls with sometimes limited and incomplete information. It provides a faster response and advice for care home staff allowing staff and clinicians to work swiftly together on resident presentations, facilitating early identification of health problems.

HealthCall’s pilot area was County Durham and Darlington, a mixed rural/urban area in the North East of England. We aim to evaluate the effectiveness of the Health Call Digital Care Homes app by looking for changes in the utilisation of unplanned secondary care as well as associated costs to service providers for the group of care home residents before and after HealthCall is implemented in each of their care homes. We do this using a large, linked dataset of healthcare interactions within County Durham and Darlington NHS Foundation Trust (CDDFT) and data from the HealthCall app.

## Methods

### Data used in the Study

We utilised data from the HealthCall Digital Care Homes app from the beginning of its rollout in December 2018 until August 2021. Three care home datasets from HealthCall covering resident enrolment, home enrolment and uploads on the app are linked to six additional routinely collected datasets from County Durham and Darlington NHS Foundation Trust hospitals (CDDFT), including ED data, inpatient data, outpatient data and community nursing data. An additional dataset containing information on patients’ hospital discharges was also used. Primary care and ambulance service data are not included.

### Linkage and Cohort Selection Criteria

Each of the datasets used in the analysis uses a pseudonymised NHS number as an individual identifier, meaning the same individual can be identified across all of the datasets. We defined the study cohort using registration data from the HealthCall app. This contains the dates when a care home resident was ‘activated’ and ‘deactivated’ from the system and the care home where the resident lives. The activation date refers to the date when a resident was added onto the HealthCall system, through implementation in the home or when they first move in afterwards. A resident may be deactivated after they die or move to a different care home.

A resident is included in the study cohort from the first date at which an observation from any of the datasets places them in the home that they were activated in. From this date they are a ‘non-HealthCall care home resident’ before their activation date. If they have no deactivation (or death) date)they are assumed to still be living in a home using HealthCall at the end of the study period. Residents are removed from the cohort when there is a ‘deactivation date’ or an identified death date for the resident in any of the datasets. A typical resident timeline is shown in figure 1. We also use observations of residents in the care homes to identify a small number of residents who were observed to be in the care homes that used the app, but were never ‘activated’ on the system who stayed as non-HealthCall residents. All residents identified were included in the analysis.

**Figure 1.**
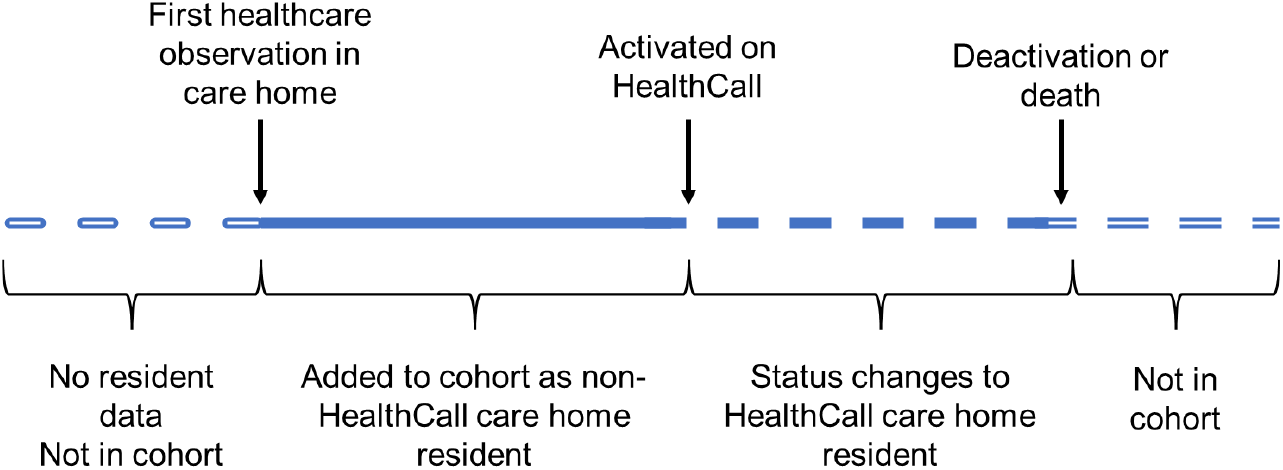
Diagram demonstrating residents’ transitions into the cohort and subsequent activation as a HealthCall Resident.

### Primary Investigation

We investigated four co-primary outcomes as potential indicators of change in unplanned secondary care usage, as well as changes in service costs associated with the introduction of HealthCall.

Service costs related to ambulance journeys to emergency departments, attendance at emergency departments, emergency inpatient stays and outpatient attendances were assigned a unit cost. These were summed to produce a total cost, at 2019/20 price levels, for each patient each month they were part of the study cohort.

Emergency department and outpatient activity were costed using their associated healthcare resource group (HRG) and National Reference costs for 2019/20 [6]. For inpatient stays, National Reference costs for 2017/18 were used [7] and inflated to 2019/20 price levels using the NHS Cost Inflation Index [8]; these costs represent the most recent for which a cost per day can be derived. Visits to care homes by healthcare professionals were costed as either by a district nurse or community matron, with one hour of time being assigned to in-person visits and 15 minutes for other types of visit [8]. The full set of unit costs are shown in Table S1 (Supplementary Materials).

### Statistical Modelling

We fitted a statistical model to each of the outcomes under investigation to understand typical patterns in these outcomes over time, and assess the impact that the introduction of HealthCall has on the expected outcomes. The gradual rollout of HealthCall over the study period as well as the occurrence of the COVID-19 pandemic during the study period means typical intervention evaluation methods such as a difference-in-difference analysis or a before and after study would not suffice.

To incorporate the different HealthCall ‘activation’ times for each of the residents we fitted resident-level Generalised Linear Mixture Models (GLMM), meaning we fit each outcome for each resident individually. We create baseline models that do not include HealthCall as a predictor variable and compare them to models which include a binary variable indicating whether a resident is activated on the HealthCall system, allowing us to determine whether HealthCall makes a significant contribution to explaining the variability of these outcomes. These models were fitted using the lme4 package in R [9].

### Baseline Model

The baseline model was a GLMM fit to resident-level outcomes, without accounting for HealthCall. This model included a random intercept for care homes, and a nested random intercept for each resident. This structure allowed for variations between care homes and residents and reflects the fact that each resident resides in only one care home. Each of the four outcome variables is a count of events of each outcome per month for each individual in the study. We use a Poisson model specification with a log-link function for the four patient outcome models.

The baseline GLMM contained five fixed effects variables, to account for typical seasonal patterns in outcomes as well as the impact of the COVID-19 pandemic, which occurred during the study period. The variables were a yearly harmonic pair (two sinusoidal curves to model cyclic fluctuations over the course of a year); month number (number of months passed since the start of the study period); monthly CDDFT COVID-19 bed days (a proxy for local COVID-19 prevalence to account for the impact of the pandemic); and pandemic wave (categorical variable to account for fluctuations in impact over the course of the pandemic). A mathematical description of the baseline model can be found in the supplementary materials.

For the economic outcome measure, costs were analysed in a similar fashion, but a two-part ‘hurdle’ model specification is used given the nature and skew of these data; the best model based on a Cullen and Frey plot adopts logistic and gamma link-functions [10]. The logistic regression estimates the probability that a resident has zero costs in a given month, while the gamma regression estimates the costs contingent on a resident having non-zero costs. Cost per resident is then calculated based on the predictions of the two regressions. This was implemented using the glmmTMB package in R.

### Testing HealthCall Effect (Step and Ongoing)

We modelled the impact of HealthCall as both an immediate step effect (binary main effect in the model) and an additional ongoing effect (as an interaction term between the binary HealthCall variable and the linear month number variable). We conducted likelihood ratio tests to assess the impact of including the step (baseline vs step model), then additionally the ongoing effect in the model (step model vs step and interaction term model).

Due to the analysis of costs requiring a two-part model, step and trend impacts of HealthCall were assessed for each of the associated regressions; the probability that a resident has zero costs (as estimated by the logistic regression) and the cost per resident for those having non-zero costs (as estimated by the gamma model).

## Results

A total of 8,702 care home residents were identified and added to the cohort. The cohort selection criteria meant that the cohort grew over time as residents were identified based on appearances in observational datasets. The overall cohort size and number of residents in each group can be seen in Figure 2. The relative size of the group of non-HealthCall residents depletes as residents are registered on HealthCall over time. Eventually almost all of the residents are registered on the system.

**Figure 2.**
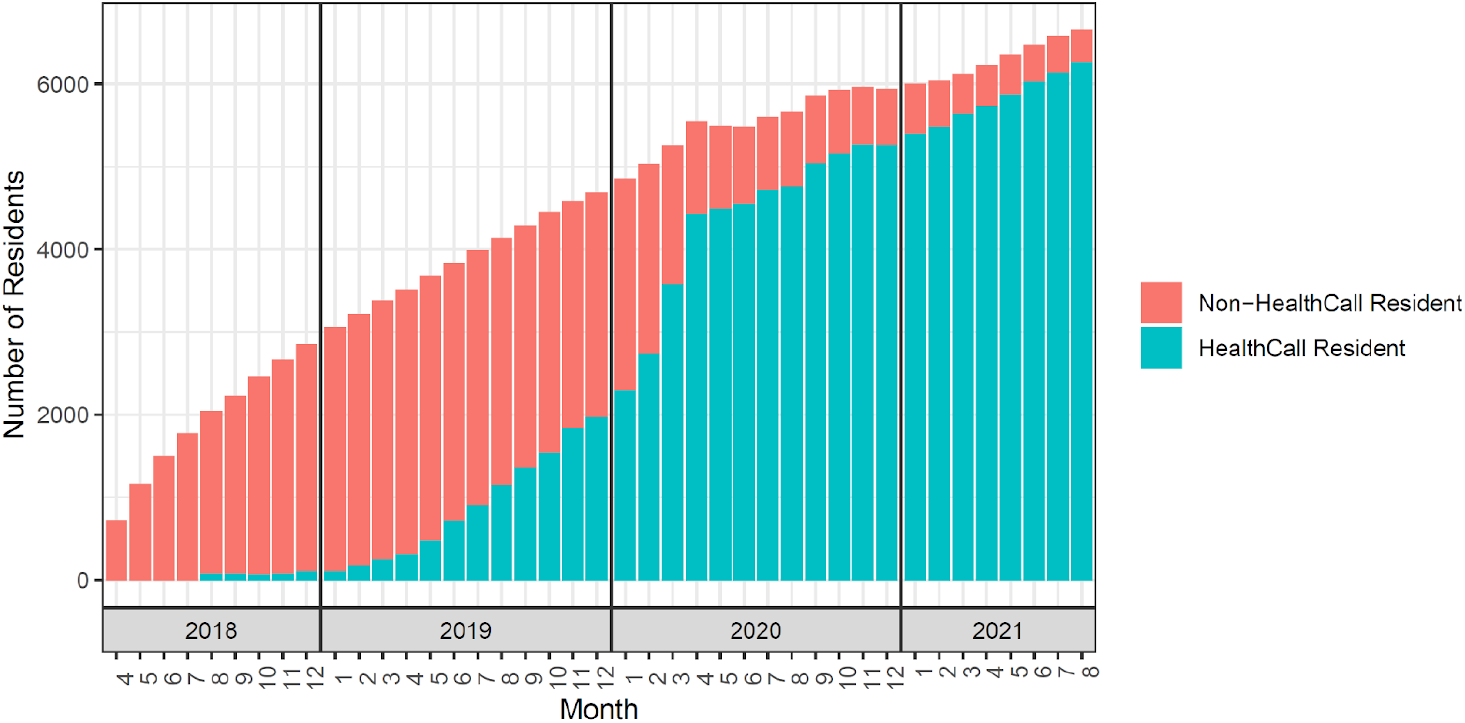
Number of residents in the cohort in each month of the study. The colours separate the groups of non-HealthCall and HealthCall residents, and the faded bars represent residents who have been removed from the cohort when deactivated or died.

A summary of the characteristics of the cohort can be found in Table 2. Of the 8,702 residents died within the study period. Some residents were deactivated from the HealthCall system for other reasons, for example, if they moved to a non-HealthCall home.

**Table 1:** Outcomes investigated in the study along with alternative hypotheses.

| Patient Outcomes | Alternative Hypothesis and Rationale |
| --- | --- |
| Monthly Emergency Attendances | Change in number of emergency attendances after introduction of HealthCall. Possible impact on potentially avoidable ED attendances. |
| Monthly Emergency Admissions | Change in number of emergency admissions after introduction of HealthCall. Possibility of improving overall resident health through proactive approach, leading to fewer emergencies. |
| Monthly number of readmissions within 28 days | Change in number of inpatient readmissions for residents after the introduction of HealthCall. Access to clinical expertise in the home may mean that residents can recover better from hospital stays, improved decision making in the hospital due to clinicians having access to more information about the resident's condition may both lead to reduced readmissions. |
| Length of stay of emergency admissions | Change in inpatient length of stay after the introduction of HealthCall. Possible early discharges due to clinicians being more confident discharging to a home that monitors residents closely leading to reduced length of stay. |
| **Economic Outcome** |  |
| Service costs | Change in the costs to the service providers due to a change in care provision after the introduction of HealthCall. |

**Table 2:** Characteristics of the cohort of care home residents included in the study. * We do not have age information for 1,394 of the residents. ** We could not calculate a Charlson Comorbidity Index for 4,671 residents due to them not having registered ICD-10 codes from their inpatient stay. Percentages are of those calculated.

|  | Median |  | IQR |  |
| --- | --- | --- | --- | --- |
| Age * | 85 |  | 79-90 |  |
| Number of Observations | 58 |  | 29-109 |  |
| Months in cohort | 19 |  | 11-31 |  |
| Months as non-HealthCall resident | 13 |  | 5-19 |  |
| Months as HealthCall resident | 14 |  | 6-18 |  |
|  | Male |  | Female |  |
| Gender | 3,086 (35%) |  | 5,616 (65%) |  |
|  | True |  | False |  |
| Died (within the study period) | 2,549 (29%) |  | 6,153 (71%) |  |
|  | 0 | 1-2 | 3-4 | ≥5 |
| Charlson Comorbidity Index ** | 324 (8%) | 2,111 (52%) | 1,292 (32%) | 324 (8%) |

Three models were fitted to each of the defined outcomes: the baseline, the HealthCall main effect (step) model and the HealthCall main effect and linear temporal interaction (ongoing) model. A demonstration of the model including the HealthCall binary variable for monthly attendances fitted over the study period can be seen in Figure 3; this shows how the model varies over time and highlights the step change between the residents on the HealthCall system (blue) and those that aren’t (red). An ongoing change was not included in this model since it was not found to be significant, hence the parallel lines. Results of the likelihood ratio tests (LRT), and the associated relative risks (derived from the coefficients) can be found in Table 3.

**Table 3:** Results table showing estimated relative risk for the HealthCall step (main effect) and monthly change (linear interaction term) and associated statistics. P-values presented here are those of the likelihood ratio test (LRT) of including this variable in the model.

| Outcome | Effect | Estimate (RR) | 95% CI | LRT p-value |
| --- | --- | --- | --- | --- |
| Emergency Attendances | Step | 0.892 | 0.846-0.941 | <0.001 |
|  | Ongoing | 1.003 | 0.999-1.008 | 0.1923 |
| Emergency Admissions | Step | 0.751 | 0.708-0.795 | <0.001 |
|  | Ongoing | 1.015 | 1.009-1.021 | 1.000 |
| 28-day Readmissions | Step | 0.709 | 0.665-0.755 | <0.001 |
|  | Ongoing | 0.982 | 0.979-0.985 | 1.000 |
| Length of Stay | Step | 0.892 | 0.817-0.974 | <0.001 |
|  | Ongoing | 0.719 | 0.665-0.755 | <0.001 |

**Figure 3.**
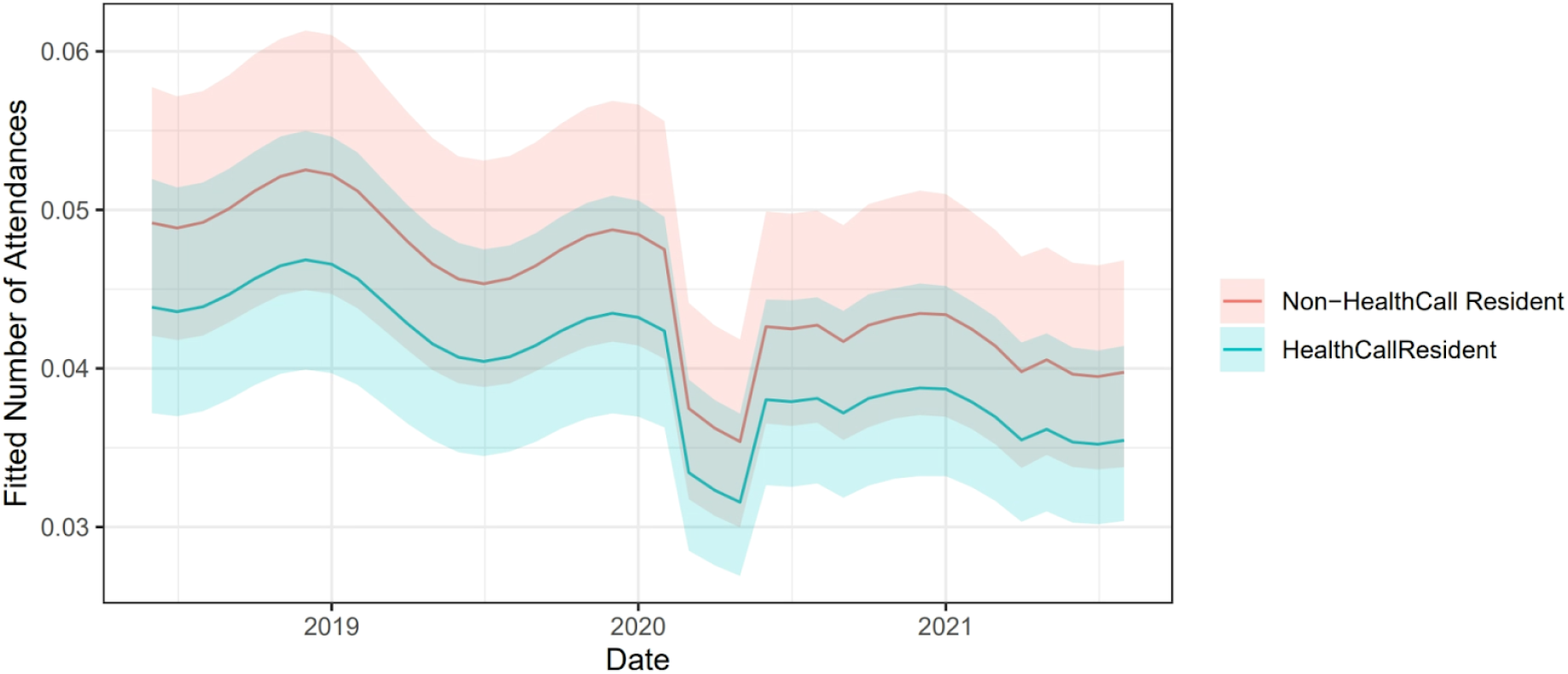
The expected number of ED attendances from the model over the study period for residents on the HealthCall system and residents not on the HealthCall system. Ribbons show 95% prediction intervals.

The number of emergency department attendances and admissions for residents on the HealthCall system were typically 11% and 25% (RR 0.89 & RR 0.75)) less than the non-HealthCall residents. For HealthCall residents 28-day readmissions were reduced by 29% and length of emergency inpatient stays were reduced by 11%, with a slope indicating decreasing length of stay for HealthCall residents of each month of the study reducing by 28% respective to the previous.

The introduction of HealthCall is estimated to produce an immediate 27% reduction in the odds of a resident-month incurring zero costs (OR=0.73), however, there is an estimated long-term trend of increasing odds per-month of zero-cost resident-months of 3% (Table 4). For non-zero cost months, HealthCall produces an immediate 24% reduction in costs. The longer-term trend in non-zero costs, while statistically significant, is small 0.03% (Table 4).

**Table 4:**
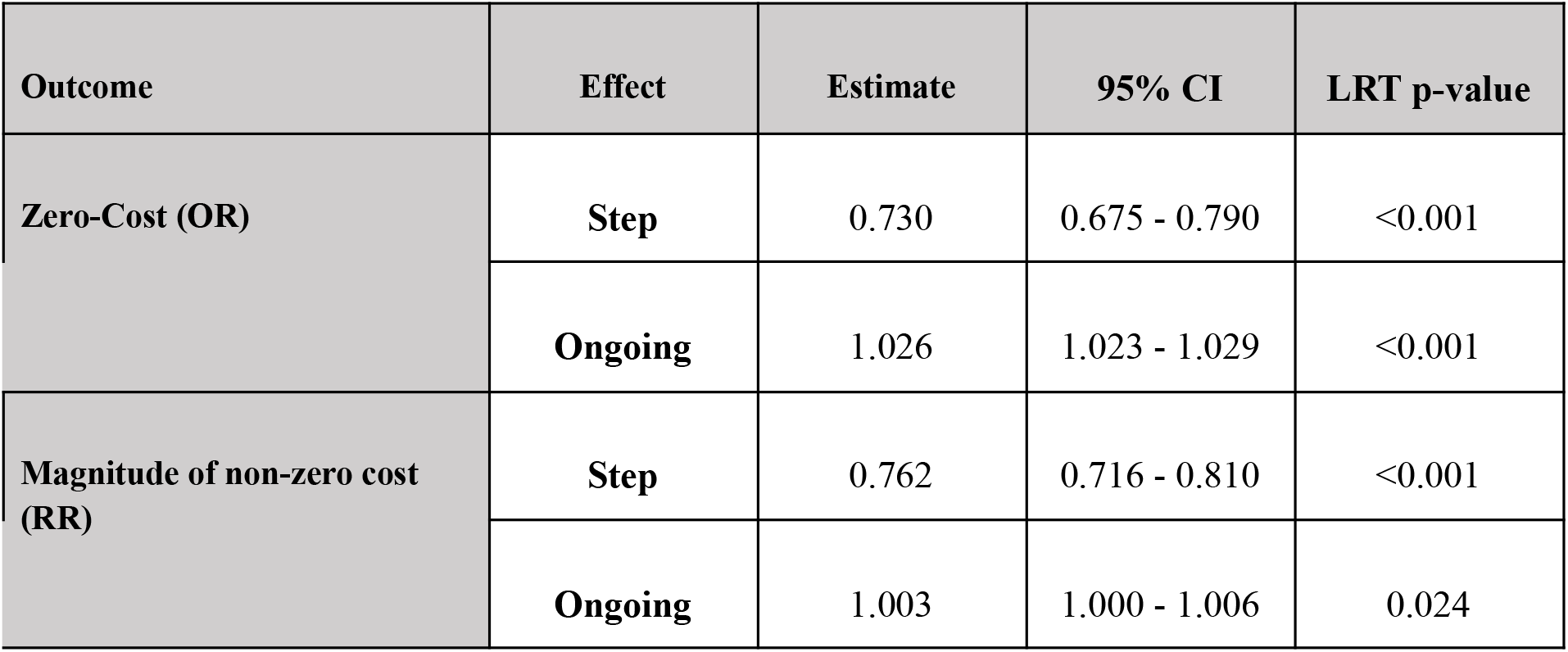
Impact of HealthCall on non-zero costs and the probability of zero monthly costs in the form of odds ratio (OR) for zero cost models and relative risks for the magnitude of costs model (RR). P-values presented here are those of the likelihood ratio test (LRT) of including this variable in the respective model.

| Outcome | Effect | Estimate | 95% CI | LRT p-value |
| --- | --- | --- | --- | --- |
| Zero-Cost (OR) | Step | 0.730 | 0.675 - 0.790 | <0.001 |
|  | Ongoing | 1.026 | 1.023 - 1.029 | <0.001 |
| Magnitude of non-zero cost (RR) | Step | 0.762 | 0.716 - 0.810 | <0.001 |
|  | Ongoing | 1.003 | 1.000 - 1.006 | 0.024 |

When combined, these predictions show that while there is an immediate decrease in the probability of a zero-cost, there is also an immediate reduction in non-zero costs, with the magnitude of the decreased costs becoming greater over time (Figure 4). The predicted values for each component part of the two-part model are shown in Figures S1 and S2. Predicted monthly costs per-resident for the four calendar years are shown in Table S3, and show a £57 reduction in cost per resident in 2018, increasing to a £113 reduction in 2021.

**Figure 4.**
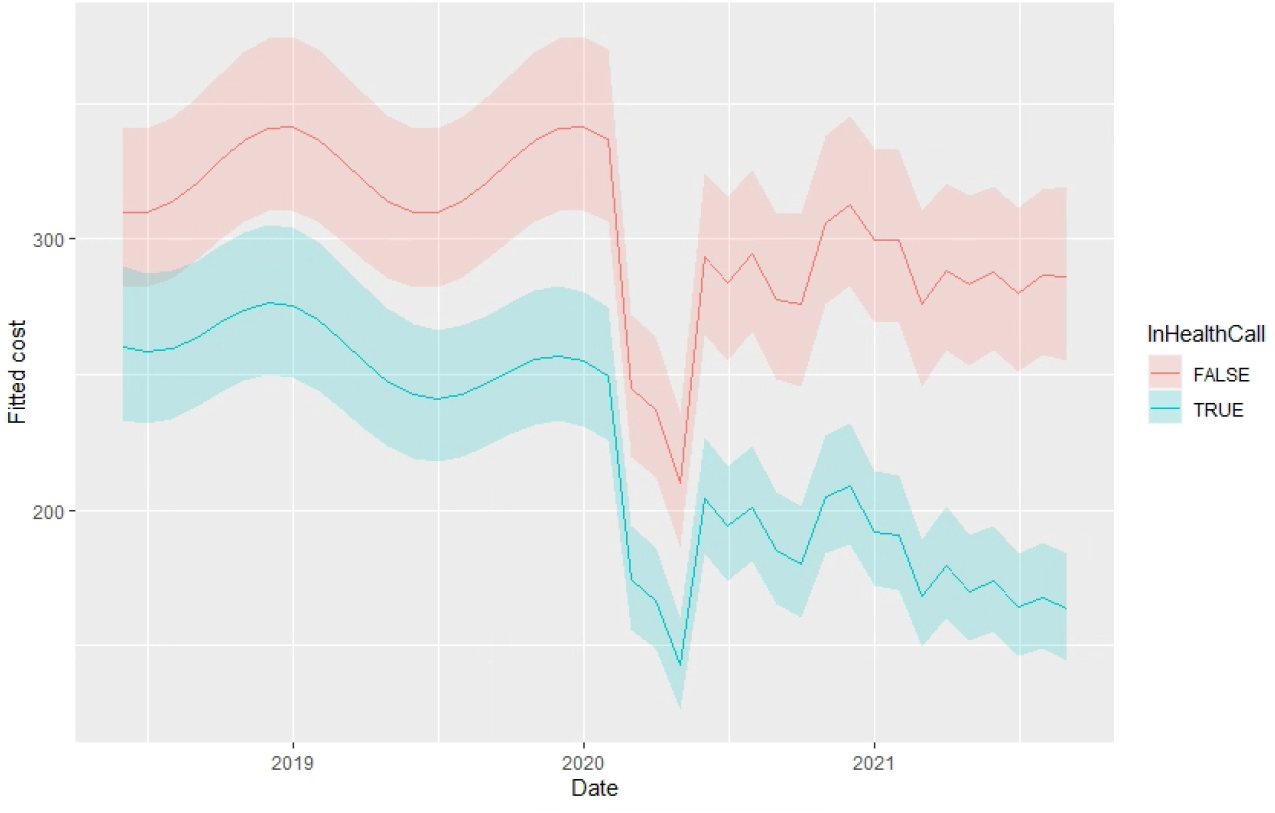
Predicted mean cost per resident of HealthCall and non-HealthCall homes over the study period.

## Discussion

### Main Findings

This study suggests that the introduction of a digital technology intervention such as HealthCall significantly reduces contacts with and costs resulting from emergency care service use by care home residents. We found that there was an 11% reduction in monthly number of emergency department attendances experienced by HealthCall registered residents compared to non HealthCall registered residents. In addition, there was a 25% reduction in emergency hospital admissions and 29% reduction in 28-day hospital readmissions for residents on the HealthCall system further reducing impact on the hospital system. HealthCall residents also experience 11% shorter emergency hospital stays, with an increasing reduction in stay over the study period of 29% compounded each month. These impacts could be due to the increase in clinical advice available to the care home staff reducing avoidable admissions, or be a consequence of the residents receiving more timely care and hence being less likely to require emergency treatment. Upskilling of staff and increasing staff confidence in the care that they give, has also been cited as an associated benefit of the HealthCall technology, and could contribute to this reduction in emergency care usage [5].

The cost analysis shows reduced health care costs for residents registered on the HealthCall system, with the magnitude of this reduction increasing over time. This trend appears mainly driven by any given resident having an increasing probability of having zero costs over time. In the first year of operation, cost savings of £57 per resident-month were estimated, which equates to £247 million for the first year across England, based on a care home population of 360,792 [4].

### Existing Literature

As part of the NHS Long Term Plan there was a promise to roll out Enhanced Health in Care Homes (EHCH) which highlights the use of technology for telehealth, remote monitoring and sharing of information to reduce uncoordinated care [11]. The HealthCall System falls within this scope and this study demonstrates the impact of the technology on the healthcare system.

In a 2016 report, The Health Foundation stated that emergency department trips could be avoided by more data sharing between care homes and NHS services and use of clinical input in care homes [2]. Our results indicate that monitoring and administering of healthcare facilitated by the HealthCall system could help address these issues. The report also highlights the challenges in accessing routinely collected data on care home residents that can be used in studies like these. The value of gathering and linking data from care homes with the wider health and social care system in order to understand patterns of service use and monitor the impact of service change must not be underestimated. Identifying ways to make routine data more available is essential in being able to evaluate interventions like these and understand their wider impact.

A number of digital interventions in care homes have been piloted in recent years, each with differing techniques to address the problems highlighted in the EHCH framework. The usage of telehealth has become particularly widespread since the start of the COVID-19 pandemic [12]. The Innovation Collaborative published a Rapid Review of remote monitoring technology in care homes [13]. The report identifies 19 remote monitoring technologies (including HealthCall) used in the UK and Ireland, with 8 case studies and one published evaluation. There is a growing body of work on telehealth initiatives for older adults outside care homes [14]. Evidence suggests that care home residents are less likely to be hospitalised and have shorter treatment times than older adults living in the community. With an ageing population, this could imply that interventions such as HealthCall can positively influence care for a wider range of older adults in the future [15]. The range of technologies becoming available highlights the need to evaluate their effectiveness using robust statistical methods, similar to those in this paper. Linkage of routinely collected hospital data with data collected in the day-to-day usage of the technologies, described here, provides a route for post-implementation evaluation of the technologies with no study-specific data collection needed.

### Limitations

The study had a number of limitations. The data contained no identifier of when a resident in the study first moved into long term care. Hospital discharge records were used to identify the date in which a resident was first observed to be in a care home. This identification method leads to a changing cohort size over time and class imbalances between the HealthCall and non-HealthCall residents. The study period was reduced prior to modelling to remove the months with the largest class imbalances. The model specification was used to account for the change in group sizes over time.

Residents were removed from the cohort when they were either deactivated from the HealthCall system or they died. Since residents have generally been activated on the HealthCall system before they are removed from the study (deactivation or death), a period of inactivity between actual and recorded deactivation could contribute to lower rates of healthcare utilisation, and therefore cost, for residents in the HealthCall group.

This study was timely, as the onset of COVID-19 during the study period led to rapid uptake of HealthCall. Our modelling aimed to disentangle the impact of HealthCall from that of the COVID-19 pandemic on healthcare utilisation, by using a proxy for COVID prevalence and a pandemic wave variable. However, the impact of the pandemic was immeasurable, results from this study may not reflect those that would have been observed during a non-pandemic period.

For the costs analysis, two notable additional weaknesses are the lack of complete ambulance service data and the nature of the community contacts data. For ambulance data, only calls resulting in an emergency department attendance have been included in our cost estimates. For community contacts, length of contact was not available, and the profession of the health care worker was poorly defined, leading to imprecise allocation of unit costs to staff. However, these issues were consistent for both HealthCall and non-HealthCall residents so confounding is likely to be minimal.

The estimated reduced costs reflect changes in the utilisation of NHS services. Not all changes across the health and social care system were included in our analysis, with the costs of the HealthCall system and associated care home activities being the most prominent of those exclusions. While the cost of HealthCall will be clear to Integrated Care Boards (ICBs) when purchasing the system, the potential costs to care homes are important to consider for successful implementation.

### Future Research

Our research provides key insights into how the introduction of a technology like HealthCall impacts healthcare utilisation and cost outcomes. Future research could investigate the HealthCall decision-making process in more detail, looking at decisions made from each individual upload from the app. This would allow for further investigation into the direct outcomes from the altered decision making provided by the app, to allow for a more detailed analysis of safety of decision-making.

The results shown in this paper are promising, but a definitive trial would help establish the true impact of the technology. Research over a larger area and longer time period, with more time before and after the intervention is introduced could improve reliability of results. The time period was limited by the data available. Randomisation of the HealthCall roll-out would be desirable to ensure findings are robust.

Replicating this research in other regions would be valuable to ascertain the generalisability of the findings. Whilst the region is diverse in terms of its mix of rural and urban areas, size/type of care homes, and size of acute hospital Trusts, it could be interpreted as being a single, regional, system. Further research could also test technologies like HealthCall in other settings such as mental health facilities.

## Data Availability

Data was collected from CDDFT and stored in a Trusted Research Environment (TRE) managed by Durham University. Informed consent was not possible as the data was anonymised. The Trust shared anonymised data after undertaking a Data Privacy Impact Assessment and a Data Transfer Agreement. Data supporting this study is not publicly available due to ethical considerations around accessing linked patient level healthcare data. The authors can no longer access the data used in this analysis. Please contact the main author for more information.

## Data Access Statement

Data was collected from CDDFT and stored in a Trusted Research Environment (TRE) managed by Durham University. Informed consent was not possible as the data was anonymised. The Trust shared anonymised data after undertaking a Data Privacy Impact Assessment and a Data Transfer Agreement. Data supporting this study is not publicly available due to ethical considerations around accessing linked patient level healthcare data. Please contact the main author for more information.

## Acknowledgements

Catherine McShane and Graham King - Facilitation of data from the County Durham and Darlington NHS Foundation Trust and the Health Call app.

Zoe Cockshott - PPIE Facilitation

PPIE Group - Helpful input

