## Supplementary Materials for "The Impact of Digital Technology in Care Homes on Unplanned Secondary Care Usage and Associated Costs"

### Supplementary Material

#### Data Description

| Healthcare Events Datasets | Description |
|---|---|
| ED | Details of attendances at 5 ED departments covered by CDDFT including the two major acute hospitals: Darlington Memorial Hospital and University Hospital of North Durham. Date and location of attendance is included, along with details of investigative procedures carried out on the patient and diagnosis codes. |
| Inpatient | Details of inpatient spells in the CDDFT hospitals. Dates for duration of overall stay and ward episodes within the stay are included. ICD-10 (International Statistical Classification of Diseases and Related Health Problems $10^{th}$ Revision) codes detailing diagnosis and comorbidities. |
| Inpatient Observations | Early Warning Scores of inpatients during their hospital stay (no constituent vital sign observations). Includes ward code of stay, date and time observation was made. |
| Outpatient | Details of outpatient appointments. Includes date and duration of interaction. Includes specialty of staff responsible for the patient. |
| Ward Episodes | Details of patient ward episodes during their hospital stays. Includes the ward code of the episode. |
| Community | Details of community appointments and callouts in the County Durham and Darlington area. Date and location type (conducted at patient's home, in community hospital etc) are included, along with care plan details indicating the reason for the interaction. |
| HealthCall | EWS observations of care home residents logged on the HealthCall app by carers. Contains the separate observations that contribute towards calculating an EWS score and the time the observations were taken. |
| **Additional Data Sets** | |
| Discharges | Summary dataset of hospital visits, including number of hospital visits and dates of discharge from hospital. Also includes care home (if applicable) of patient mined from hospital records, and date of death (if applicable) contained in hospital records of the patient. Used as a lookup table for patient death dates. |
| HealthCall Referrals | Dates of activation and deactivation of care home residents on the HealthCall system. Activation dates refer to the date they are first put onto the HealthCall system, may be when HealthCall first goes live in the care home, or when the resident first moves to the care home. Conversely, deactivation dates may refer to the date a resident leaves the care home (moves care home or goes back to own accommodation) or dies. The data identifies the most recent care home each resident has been assigned to, providing an indicator of each resident's care home. |
| HealthCall Implementation | Dates each HealthCall care home 'went live' and implemented HealthCall. This is the only non-patient level dataset. |

#### Model Specification

Let $y$ be the outcome variable of the investigation. $i$, $(i = 1...n)$ corresponds to a resident from care home $j$, $(j = 1...g)$.

Outcome $Y$ is the monthly count of events for each resident (this will vary dependent on the outcome we are modelling), we assume that these counts follow a Poisson distribution. μ is the estimated value of the distribution. $h$ is the link function between the linear predictor and outcome $y$, decided depending on the distribution of $Y$.

The proposed baseline model will take the form;

$$Y_{ijk} \sim Poisson\left(y_{ijk}; \lambda_{ijk}\right)$$

$$\eta_{ijk} = \ln ln\left(\lambda_{ijk}\right) = \beta_0 + \beta_1 x_{1ijk} + \beta_2 x_{2ijk} + \ldots + b_j + c_{ij} + \epsilon_{ijk}$$

$$b \sim Normal\left(0, \tau_b^2 I_g\right)$$

$$c_j \sim Normal\left(0, \tau_c^2 I_n\right)$$

$$\epsilon_{ijk} \sim Normal\left(0, \sigma_{ijk}\right),$$

where $\eta$ is the linear predictor, $\beta$ are the regression coefficients. $x_{pijk}$ is the $k$th observation of the $p$th variable of individual $i$ from care home $j$. $b_0$ corresponds to variation on a care home level, and $c_0$ corresponds to variation on an individual level. The individual level random intercept is nested within the care home level since each individual lives in only one care home.

(Below from economics)

*Table S1: Unit costs and sources*

| Item of resource | Unit cost (£, 2019/20) | Source |
|---|---|---|
| District nurse face-to-face visit | 49 | PSSRU, 2020 |
| District nurse other visit | 25 | PSSRU, 2020 |
| Community matron face-to-face visit | 59 | PSSRU, 2020 |
| Community matron other visit | 30 | PSSRU, 2020 |
| Ambulance conveyance to emergency department | 292 | NRC, 2019/20 |
| Emergency department attendance | Various | NRC, 2019/20 |
| Outpatient attendance | Various | NRC, 2019/20 |
| Inpatient cost per day (non-elective) | Various | NRC, 2017/18 |

*Table S3: Predicted monthly costs per resident by calendar year*

| | No HealthCall | HealthCall | Difference |
|---|---|---|---|
| 2018 | 322.96 | 265.95 | 57.01 |
| 2019 | 325.30 | 253.93 | 71.37 |
| 2020 | 284.51 | 197.24 | 87.27 |
| 2021 | 287.48 | 174.52 | 112.96 |

*Figure S1: Predicted probability of resident having zero cost*

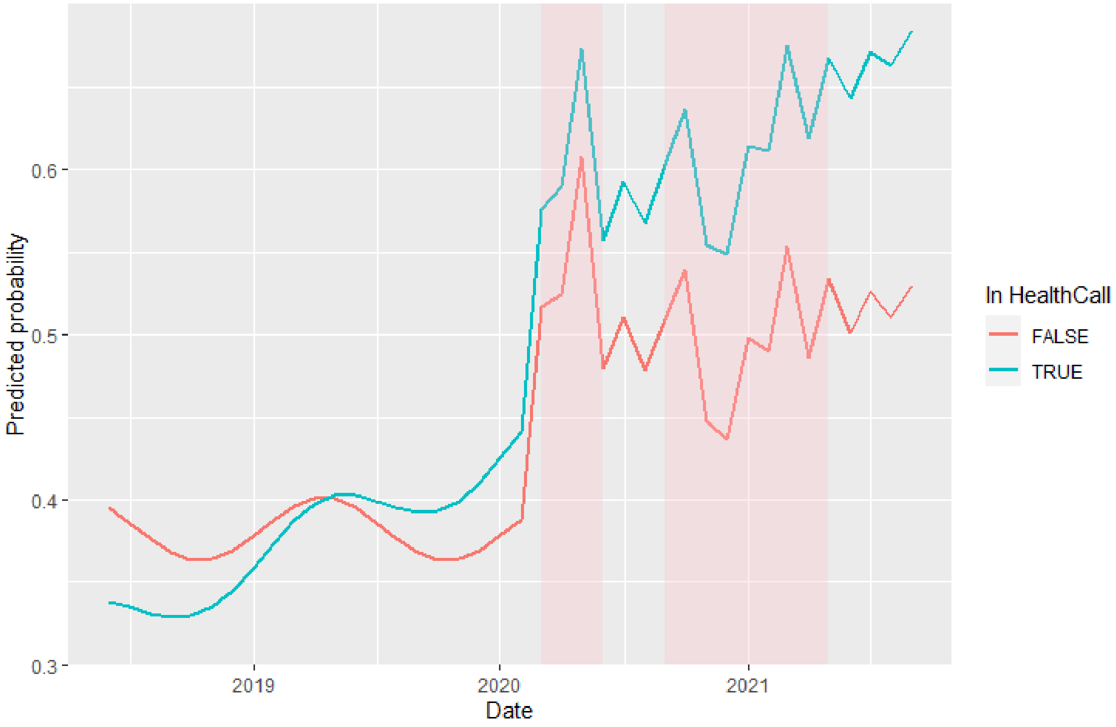


HealthCall is associated with an immediate reduction in the probability of a resident having a zero cost relative to non-HealthCall homes, as illustrated by the lower line in 2018 (OR=0.730, $p<0.001$). However, the probability of a Health Call resident having a zero cost increases monthly relative to non-HealthCall residents (OR=1.026, $p<0.001$). Consequently, the probability of a resident having zero costs becomes greater in Health Call homes.

*Figure S2: Predicted cost of residents with non-zero costs*

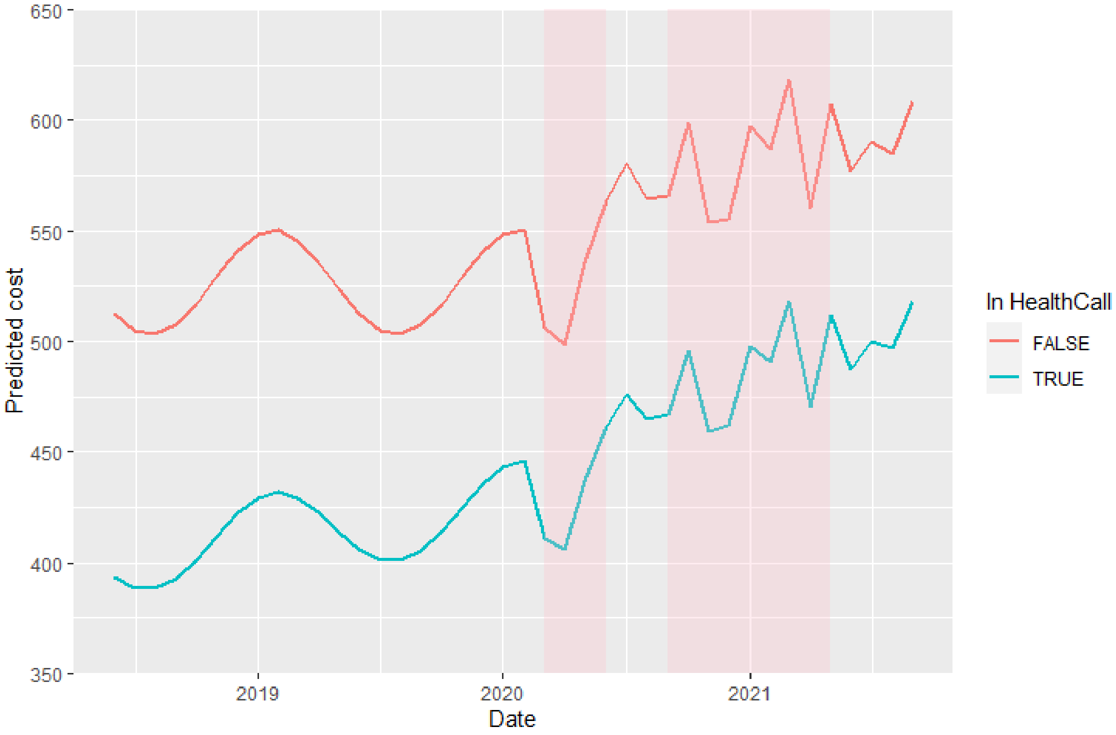


Health Call is associated with an immediate reduction in the predicted costs of residents with non-zero health care costs, relative to non-HealthCall residents (PR=0.762, p<0.001). This difference reduces marginally, over time (PR=1.003, p=0.024).
